# Assessing the level and determinants of COVID-19 Vaccine Confidence in Kenya

**DOI:** 10.1101/2021.06.11.21258775

**Authors:** Stacey Orangi, Jessie Pinchoff, Daniel Mwanga, Timothy Abuya, Mainga Hamaluba, George Warimwe, Karen Austrian, Edwine Barasa

## Abstract

The government of Kenya has launched a phased rollout of COVID-19 vaccination. A major barrier is vaccine hesitancy; the refusal or delay of accepting vaccination. This study evaluated the level and determinants of vaccine hesitancy in Kenya. We conducted a cross-sectional study administered through a phone-based survey in February 2021 in four counties of Kenya. Multivariate logistic regression was used to identify individual perceived risks and influences, context-specific factors, and vaccine-specific issues associated with COVID-19 vaccine hesitancy. COVID-19 vaccine hesitancy in Kenya was high: 60.1%. Factors associated with vaccine hesitancy included: older age, lower education level, perceived difficulty in adhering to government regulations on COVID-19 prevention, less adherence to wearing of face masks, not having ever been tested for COVID-19, no reported socio-economic loss as a result of COVID public-health restriction measures, and concerns regarding vaccine safety and effectiveness. There is a need for the prioritization of interventions to address vaccine hesitancy and improve vaccine confidence as part of the vaccine roll-out plan. These messaging and/or interventions should be holistic to include the value of other public health measures, be focused and targeted to specific groups, raise awareness on the risks of COVID-19 and effectively communicate the benefits and risks of vaccines.

## BACKGROUND

The Coronavirus disease (COVID-19) caused by the novel severe acute respiratory syndrome coronavirus-2 (SARS-CoV-2) was declared a pandemic on the 11^th^ of March 2020 with the first case in Kenya confirmed on the 12^th^. As of June 4^th^, 2021, there have been 171,658 confirmed COVID-19 cases and 3,223 deaths in Kenya [1]. To control the pandemic, non-pharmaceutical interventions (NPIs) have been put in place in Kenya, as in other settings, to slow down the transmission of the virus thus averting COVID-19 morbidity and mortality. These measures have ranged from physical distancing measures, movement restrictions, closure of schools, sanitation measures, testing, and wearing of face masks in public places. These NPIs also have potential indirect effects that are multifaceted including slow economic growth, financial hardships, reduced access to essential health services, food insecurity, gendered impacts, and widening inequality in access to education [2,3]. Given the indirect effects that these NPIs present and based on risk assessments, they have been implemented with different intensities over time in Kenya [3].

Vaccines are a key intervention in the response against the COVID-19 pandemic, with the potential to protect populations from infection, severe disease and death, and block transmission from infected to uninfected people. The development and deployment of COVID-19 vaccines have thus been prioritized [4]. As of 2^nd^ May 2021, there were about 275 vaccine candidates in clinical development, the majority of which are in the pre-clinical stages (67%) [5]. There are about 91 vaccines in clinical trials and 6 vaccines approved for emergency use. [5,6].

The government of Kenya plans to vaccinate 50% of all adult populations by end of June 2022 in a phased approach while maintaining a prioritization matrix. As a result, Kenya launched the rollout of COVID-19 vaccination procured through the COVID-19 Vaccines Global Access facility (COVAX) in March 2021 [7]. The vaccination is being rolled out progressively with the prioritized population being: 1)Essential workers (including healthcare providers, teachers, security personnel) which is an estimated 1.25 million Kenyans 2) Individuals at risk of severe disease including older adults (58 years and above) and those above 18 years with co-morbidities – which targets 9.76 million Kenyans 3) Individuals at high risk of infection such as people 18 years and above in congregate settings, as well as the hospitality and tourism industry – targeting 4.9 million Kenyans [7]. As of June 4^th^, 2021, there were 969,561 COVID-19 vaccine doses administered in Kenya [1].

Evidence suggests that for herd immunity to be attained, about 60-70% of the population would need to receive the COVID-19 vaccine [8,9]. However, one major barrier to achieving high levels of vaccine coverage is vaccine hesitancy; defined as the refusal or delay in acceptance of vaccines, despite their availability [10]. It lies across a spectrum between total acceptance and total refusal [10]. In sub-Saharan Africa, studies suggests that some of the reasons of COVID-19 vaccine hesitancy include, negative perceptions of the pharmaceutical industries, concerns on vaccine safety and/or the source of the vaccine, lack of confidence in the government, and vaccine costs [11–14]. Vaccine hesitancy is however context-specific and varies across time and place [15]. Kenya reports a high vaccine confidence against childhood diseases, with 89% reporting vaccines as safe, 87% reporting them as being effective, and 97% perceived importance for childhood vaccination [16]. However, there is limited evidence and understanding of the public willingness to accept; and the confidence they place on the COVID-19 vaccine in Kenya, which mainly targets the adult population. This study, therefore, aims to determine the level of COVID-19 vaccine hesitancy in Kenya and report on its determinants.

## METHODS

### Study design

This study employed a cross-sectional study design to administer a knowledge, attitudes, and practices survey through a phone-based platform. The survey was conducted in February 2021.

### Study population and setting

The survey was administered to participants sampled from households in four existing Population Council prospective cohort studies across four counties; Nairobi, Wajir, Kilifi, and Kisumu [17].

Specifically, in Nairobi County the target population was from five urban informal settlements: 2,565 households in Huruma and Kibera enrolled in the Adolescent Girls Initiative-Kenya (AGI-K) study [17,18] and 4,519 households in Dandora, Kariobangi, and Mathare enrolled in the NISITU program study [17,19]. In Wajir County, an arid region in north-eastern Kenya, the target population were households from 79 villages in Wajir East, Wajir West, and Wajir South sub-counties enrolled in the AGI-K study [17,18]. In Kilifi County, a coastal region in Kenya, households from three sub-counties (Ganze, Kaloleni, and Magarini) enrolled in the Nia Project formed the target population [20]. Lastly in Kisumu County, located in Western Kenya, the target populations were households in Nyalenda area (one of the largest informal settlements in Kisumu) and Kolwa East (a peri-urban area) who were enrolled in the PEPFAR DREAMS study cohort [17]. This is illustrated in Table 1.

**Table 1:** Study population and sample size.

| County | Location | Underlying Population Council cohort | Sample size |
| --- | --- | --- | --- |
| Nairobi | 5 Informal Settlements (Kibera, Huruma, Dandora, Kariobangi, Mathare) | Adolescent Girls Initiative-Kenya (AGI-K) study<br>NISITU Program | n=1,117 (697 females) |
| Wajir | 79 villages in 3 sub-counties (Wajir East, Wajir West, Wajir South) | Adolescent Girls Initiative-Kenya (AGI-K) study | n=1,218 (833 females) |
| Kilifi | 3 sub-counties (Ganze, Kaloleni, Magarini) | Nia Project | n=1,096 (657 females) |
| Kisumu | 1 informal settlement (Nyalenda) &<br>1 peri-urban area (Kolwa East) | • PEPFAR DREAMS study | n=704<br>(593 females) |

### Sample size and sampling procedure

Households with available phone numbers were randomly sampled from the four existing cohorts using a ratio of 1:3 for male to female interviews. Due to the nature of the sampling frame described above, the randomly sampled participants were from households with at least one adolescent. Households that solely constituted adult residents or adults and very young children only were not eligible for inclusion in the initial cohorts and therefore not represented in this study.

### Data collection tool

The data collection tool was a knowledge, attitudes, and practices survey that collected information on 1) socio-demographic background information 2) the knowledge, attitudes, and practices reported by households concerning COVID-19 3) the barriers to adoption of non-pharmaceutical interventions for COVID-19 prevention 4) the social, economic, education and health effects of COVID-19 prevention measures on adults 5) the level and determinants of vaccine hesitancy. The latter being the focus of this study.

The data collection tool was designed using validated measures where possible such as the WHO SAGE vaccine hesitancy tool, existing COVID-19 vaccine hesitancy tools [21,22], and was also informed by local Kenyan researchers. Questions on the determinants of vaccine hesitancy were categorized into three broad groups adapted from the WHO SAGE vaccine hesitancy tool [21]. The first category reported on individual and group influences. These included questions on the individual’s perceived risk of getting COVID, the ease of following government regulations, societal perception of having COVID-19, the individual adherence to wearing masks and having ever been tested for COVID, and the socio-economic impact of COVID-19 on the individual. The second category of questions on determinants of vaccine hesitancy focused on the context; specifically, looking into the trusted sources of information for COVID and the perceived level of community support for COVID-19 prevention measures. Lastly, the third category included vaccine-specific questions such as concerns on the COVID-19 vaccine side-effects and effectiveness, access to the vaccination site, fear of needles, being too busy to be vaccinated, and religious and cultural reasons for refusing vaccination.

The tools were in English and translated to Swahili, Dholuo and Somali, piloted, and administered by local interviewers. Data was collected using Open Data Kit.

### Ethical Approval

Ethical approval for this study was obtained from both Population Council Institutional Review Board (p936) and AMREF Ethics and Scientific Review Committee (P803/2020). Before data collection, verbal informed consent was obtained from all participants 18 years and over. Participants were told they could terminate the survey at any time or refuse to answer specific questions. Participants were informed beforehand that they would be reimbursed 100 Kenyan shillings (~US$1) for their time (transferred via M-PESA mobile money).

### Data Analysis

The socio-demographic characteristics of the sample population were described by computing descriptive statistics. A cross-tabulation analysis was performed to determine the level of vaccine hesitancy among the respondents’ sociodemographic characteristics using chi-squared tests.

Multivariate logistic regression analyses, accounting for the counties as clusters, were performed to compute the adjusted odds ratio (aOR) with a 95% confidence interval. Vaccine hesitancy was the dependent variable and was dichotomized as either accepting (i.e. very likely to get the vaccine) or hesitant (showing some level of hesitancy i.e. somewhat likely, somewhat unlikely, or very unlikely to get the vaccine). Socio-demographic characteristics, individual risks and perceptions, contextual factors, and vaccine-specific issues were included as predictor variables for vaccine hesitancy. The description of the dependent and independent variables is illustrated in supplementary table 1. Predictor variables were included in the multivariate model if found to be significant at a 0.05 significance level in the crude logistic regression. Multicollinearity of the variables was assessed using variance inflation factors and the Hosmer-Lemeshow goodness-of-fit test was used to ensure that the model adequately fit the data. The significance level was set at <0.05 and Stata version 15.0 (Stata Corporation, College Station, Texas, USA) was used for the data analyses.

## RESULTS

### Descriptive statistics analyses

The socio-demographic characteristics of the 4,136 respondents who participated in this study are shown in Table 2. The mean age of the respondents was 40.8 years (SD 12.6) and the average household size of the respondents was 7.5 (SD 4.4). Most of the respondents were female (67.2%), residents of rural counties (56.0%), married (72.7%), had either no schooling or primary school level education) as the highest education level (70.7%), and were from the lowest wealth tertile (36.7%).

**Table 2:**
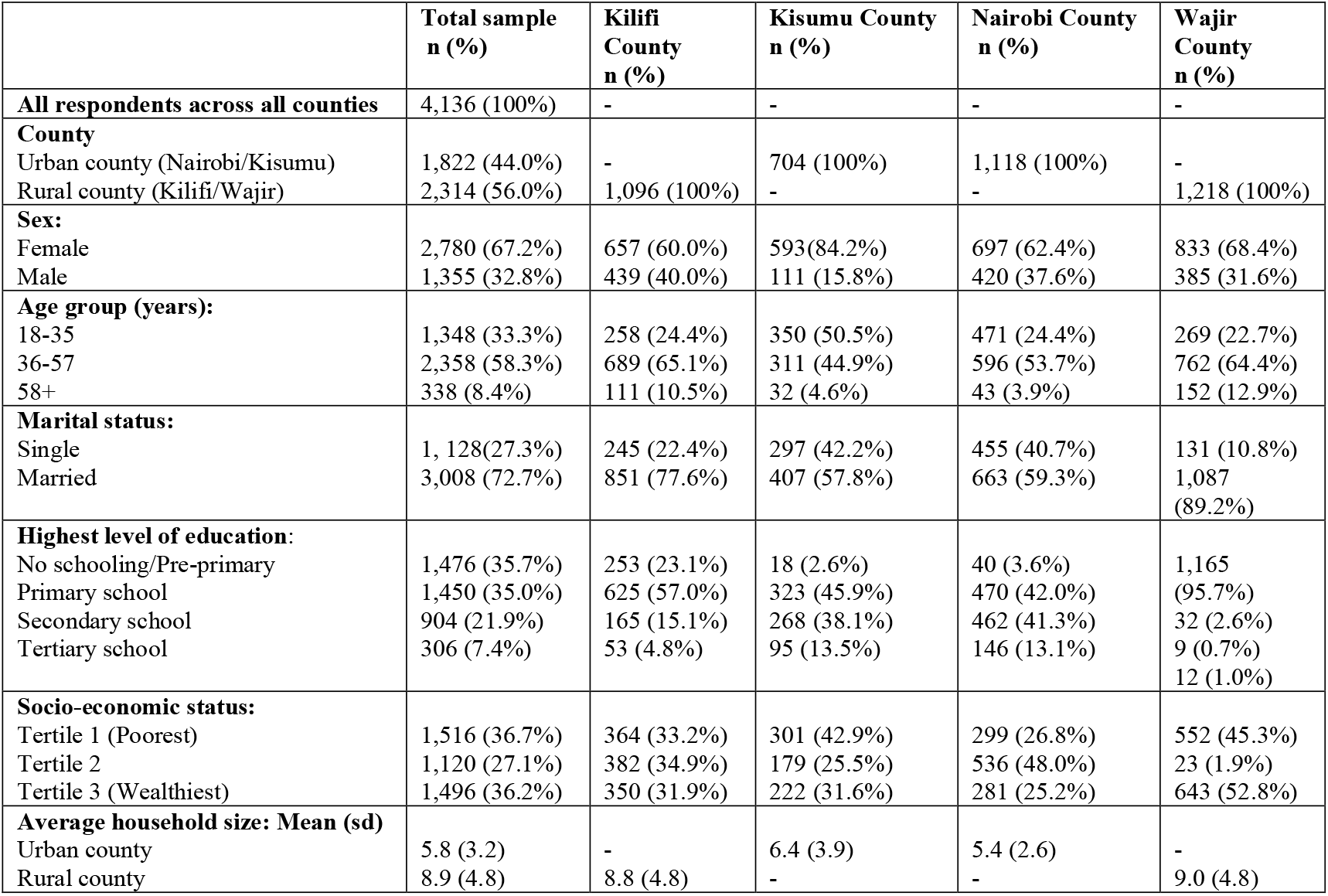
Socio-demographic characteristics by county.

Among the respondents, the amount participants were willing to pay for the vaccine (if it was not available for free) was reported at USD 3.23 (KES 323.06 (SD 1407.69)) and the overall reported hesitancy towards the COVID-19 vaccine was 60.6% (n=2,507). The overall level of vaccine hesitancy and across the counties is illustrated in figure 1.

**Figure 1:**
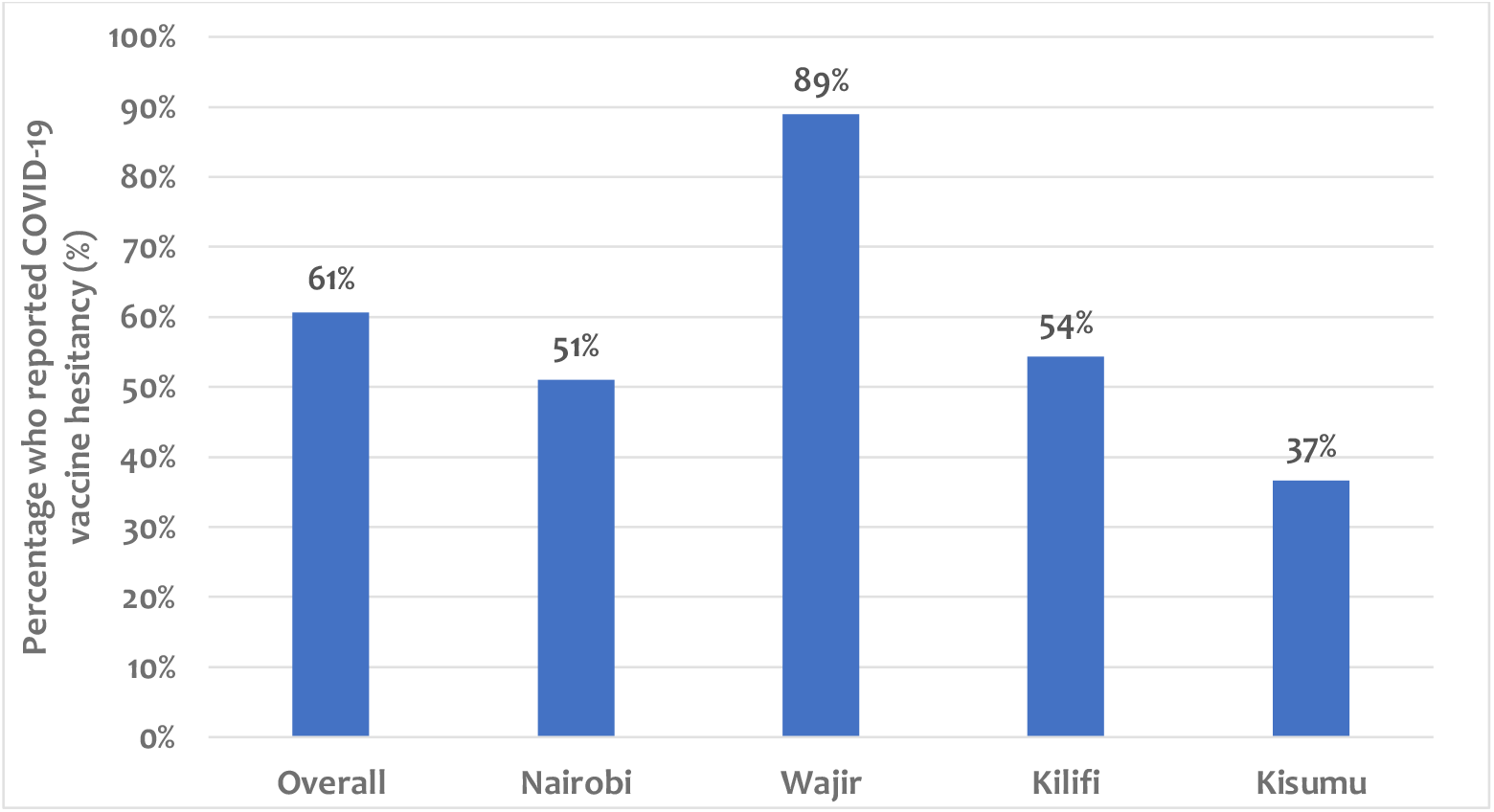
Level of Vaccine Hesitancy Across Study Counties.

### Bivariate analysis

Table 3 shows bivariate associations between socio-demographic characteristics and the level of COVID-19 vaccine hesitancy in Kenya. Across all study counties, of the 338 respondents over 58 years, 64.5% (n=218) reported vaccine hesitancy. For those who were married (n=3,008), 62.3% (n=1,875) were likely to report vaccine hesitancy. Of the 1,476 respondents who had no schooling or only pre-primary level of education, 1,201 (81.4%) reported vaccine hesitancy.

**Table 3:** Bivariate associations between socio-demographic factors and intent of COVID-19 vaccine uptake among respondents in study counties.

|  | Overall |  | Kilifi County |  | Kisumu County |  | Nairobi County |  | Wajir County |  |
| --- | --- | --- | --- | --- | --- | --- | --- | --- | --- | --- |
|  | Vaccine hesitant individuals n (%) | p-value | Vaccine hesitant individuals n (%) | p-value | Vaccine hesitant individuals n (%) | p-value | Vaccine hesitant individuals n (%) | p-value | Vaccine hesitant individuals n (%) | p-value |
| **All respondents** | 2,507 (60.6%) | - | 595 (54.3%) | - | 258 (36.7%) |  | 571 (51.1%) |  | 1,083 (88.9%) |  |
| **County** |  |  |  |  |  |  |  |  |  |  |
| Urban county (Nairobi/Kisumu) | 829 (45.1%) | **0.000** |  |  |  |  |  |  |  |  |
| Rural county (Kilifi/Wajir) | 1,678 (72.5%) |  |  |  |  |  |  |  |  |  |
| **Sex:** |  |  |  |  |  |  |  |  |  |  |
| Female | 1,689 (60.8%) | 0.776 | 383 (58.3%) | **0.001** | 214 (36.1%) | 0.476 | 355 (50.9%) | 0.933 | 737 (88.5%) | 0.471 |
| Male | 817 (60.3%) |  | 212 (48.3%) |  | 44 (39.6%) |  | 215 (51.2%) |  | 346 (89.9%) |  |
| **Age group (years):** |  |  |  |  |  |  |  |  |  |  |
| 18-35 | 766 (56.8%) | **0.002** | 137 (53.1%) | 0.423 | 139 (39.7%) | **0.003** | 255 (54.1%) | 0.177 | 235 (87.4%) | 0.593 |
| 36-57 | 1,467 (62.2%) |  | 384 (55.7%) |  | 113 (36.3%) |  | 289 (48.5%) |  | 681 (89.4%) |  |
| 58+ | 218 (64.5%) |  | 55 (49.6%) |  | 3 (9.4%) |  | 23 (53.5%) |  | 137 (90.1%) |  |
| **Marital status:** |  |  |  |  |  |  |  |  |  |  |
| Single | 632 (56.0%) | **0.000** | 147 (60.0%) | **0.042** | 113 (38.1%) | 0.510 | 254 (55.8%) | **0.008** | 118 (90.1%) | 0.654 |
| Married | 1,875 (62.3%) |  | 448 (52.6%) |  | 145 (35.6%) |  | 317 (47.8%) |  | 965 (88.8%) |  |
| **Highest level of education:** |  |  |  |  |  |  |  |  |  |  |
| No schooling/Pre-primary | 1,201 (81.4%) | **0.000** | 144 (56.9%) | 0.649 | 6 (33.3%) | **0.005** | 15 (37.5%) | **0.001** | 1,036 (88.9%) | 0.724 |
| Primary school | 673 (46.4%) |  | 329 (52.6%) |  | 100 (31.0%) |  | 215 (45.7%) |  | 29 (90.6%) |  |
| Secondary school | 456 (50.4%) |  | 92 (55.8%) |  | 104 (38.8%) |  | 253 (54.8%) |  | 7 (77.8%) |  |
| Tertiary school | 177 (57.8%) |  | 30 (56.6%) |  | 48 (50.5%) |  | 88 (60.3%) |  | 11 (91.7%) |  |
| **Socio-economic status:** |  |  |  |  |  |  |  |  |  |  |
| Tertile 1 (Poorest) | 949 (62.6%) | **0.000** | 195 (53.6%) | 0.657 | 108 (35.9%) | 0.321 | 160 (53.5%) | 0.395 | 486 (88.0%) | 0.655 |
| Tertile 2 | 547 (48.8%) |  | 203 (53.1%) |  | 60 (33.5%) |  | 263 (49.1%) |  | 21 (91.3%) |  |
| Tertile 3 (Wealthiest) | 1,011 (67.6%) |  | 197 (56.3%) |  | 90 (40.5%) |  | 148 (52.7%) |  | 576 (89.6%) |  |

**Table 4:** Logistic regression analysis for factors potentially associated with COVID-19 vaccine hesitancy among respondents in Kenya.

|  | aOR (95% CI) | p-value |
| --- | --- | --- |
| **Socio-demographic factors** |  |  |
| **County** |  |  |
| Urban county (Nairobi/Kisumu) | Ref |  |
| Rural county (Kilifi/Wajir) | 1.48 (0.99-2.21) | 0.056 |
| **Sex** |  |  |
| Female | Ref |  |
| Male | 1.10 (0.88-1.36) | 0.417 |
| **Age group (years):** |  |  |
| 18-35 | Ref |  |
| 36-57 | **1.08 (1.01-1.15)** | **0.018** |
| 58+ | 0.82 (0.52-1.29) | 0.385 |
| **Marital status:** |  |  |
| Single | Ref |  |
| Married | 0.90 (0.75-1.08) | 0.267 |
| **Education:** |  |  |
| No schooling/Pre-primary | Ref |  |
| Primary school | **0.31 (0.19-0.51)** | **0.000** |
| Secondary school | **0.41 (0.25-0.66)** | **0.000** |
| Tertiary school | **0.47 (0.32-0.71)** | **0.000** |
| **Socio-economic status:** |  |  |
| Tertile 1 (Poorest) | Ref |  |
| Tertile 2 | **0.84 (0.75-0.93)** | **0.001** |
| Tertile 3 (Wealthiest) | **1.12 (1.00-1.24)** | **0.045** |
| **Individual influences, risks, and perceptions** |  |  |
| **Perceived COVID infection risk** |  |  |
| Some risk | Ref |  |
| No risk | 1.47 (0.96-2.25) | 0.076 |
| **Perceived ability to adhere to government regulations regarding COVID-19 prevention** |  |  |
| Easy to adhere | Ref |  |
| Difficult to adhere | **1.49 (1.25-1.80)** | **0.000** |
| **Wearing of masks (now compared to when COVID began)** |  |  |
| Wear masks more or the same | Ref |  |
| Wear masks less | **1.57 (1.37-1.81)** | **0.000** |
| **Testing for COVID** |  |  |
| Ever tested for COVID | Ref |  |
| Never tested for COVID | **1.93 (1.65-2.26)** | **0.000** |
| **Socio-economic impact of COVID measures** |  |  |
| Socio-economically affected by measures | Ref |  |
| Not socio-economically affected by measures | **1.60 (1.17-2.19)** | **0.003** |
| **Context** |  |  |
| **Government and media as a trusted source of information** |  |  |
| No | Ref |  |
| Yes | 0.86 (0.61-1.23) | 0.411 |
| **Healthcare providers as a trusted source of information** |  |  |
| No | Ref |  |
| Yes | 0.88 (0.67-1.17) | 0.384 |
| **Vaccine specific issues** |  |  |
| **Vaccine side effects concerns** |  |  |
| No | Ref |  |
| Yes | **5.81 (5.01-6.74)** | **0.000** |
| **Don't think the vaccine is effective** |  |  |
| No | Ref |  |
| Yes | **1.91 (1.17-3.13)** | **0.010** |
| **Hard to access vaccination sites** |  |  |
| No | Ref |  |
| Yes | 0.96 (0.60-1.53) | 0.859 |
| **Scared of needles** |  |  |
| No | Ref |  |
| Yes | 1.26 (0.97-1.64) | 0.080 |
| **Religious and cultural reasons** |  |  |
| No | Ref |  |
| Yes | 0.69 (0.36-1.34) | 0.273 |
| **Too busy to get vaccinated** |  |  |
| No | Ref |  |
| Yes | 1.26 (0.82-1.94) | 0.298 |

### Multivariate analysis

The multivariate logistic regression analysis for socio-demographic factors, individual influences, contextual factors, and vaccine-specific factors as predictors of vaccine hesitancy among the respondents is illustrated in Table 3. In the multivariate model, respondents who were between the age of 36-57 years had 1.08 times higher odds of reporting vaccine hesitancy (aOR:1.08; 95% CI:1.01-1.15) as compared to those between 18-35 years. Further, those who had a tertiary level of education had 0.47 lower odds of being vaccine hesitant (aOR:0.47; 95% CI:0.32-0.71).

Regarding individual influences, those who reported difficulty in adhering to government regulations regarding COVID-19 prevention had 1.49 higher odds of being more likely to be vaccine hesitant (aOR:1.49; 95%CI:1.25-1.80). Similarly, those who reported wearing face masks less often now as compared to when COVID-19 started had 1.57 higher odds of being vaccine hesitant (aOR:1.57; 95% CI:1.37-1.81) as compared to those whose adherence to wearing face masks had remained the same or improved over time. Additionally, those who had previously never tested for COVID-19 had 1.93 times higher odds of reporting vaccine hesitancy (aOR:1.93; 95% CI:1.65-2.26). Individuals who were not socio-economically affected by COVID-19 measures had 1.60 higher odds of being vaccine hesitant (aOR:1.60;95% CI:1.17-2.19) as compared to those who were socio-economically affected.

In relation to vaccine-specific issues, respondents who were concerned about the COVID-19 vaccine side effects or were concerned about vaccines’ effectiveness had 5.81 higher odds (aOR:5.81; 95% CI:5.01-6.74) and 1.91 higher odds (aOR:1.91; 95%CI:1.17-3.13) to be vaccine hesitant respectively.

## DISCUSSION

This study set out to evaluate the level and determinants of COVID-19 vaccine hesitancy in Kenya. We make several reflections from the findings. First, the overall level of COVID-19 vaccine hesitancy reported is high (60.61%), as compared to childhood vaccine acceptance in Kenya: Specifically, reported perceived childhood vaccine safety, effectiveness, and relative importance of common childhood vaccines is greater than 87% in Kenya [16]. This highlights the need for the Kenyan government to prioritize interventions to address vaccine hesitancy and improve vaccine confidence as part of its vaccine roll-out plan; examine the socio-demographic factors, individual effects and perceived risks, context, and vaccine-specific issues that affect vaccine hesitancy. This level of hesitancy is consistent with findings elsewhere. For instance, a survey done across 19 countries reported 71.5% of the respondents were very likely or somewhat likely to accept an available COVID-19 vaccine that is proven safe and effective, with differences in vaccine acceptance ranging from 90% in China to 55% in Russia [23]. In sub-Saharan Africa, surveys have reported 84.6% of Cameroonians, 52% of South Africans, and 50% of Zimbabweans to be hesitant or would reject the COVID-19 vaccine [13,24,25].

Second, demographic and socio-economic factors that determine vaccine hesitancy include age, level of education, and socio-economic status. In contrast to some studies [22,26–28], we report a higher likelihood of vaccine hesitancy between those of a higher age group (36-57 years) as compared to younger people (18-35 years). However, our findings align with some settings such as Ireland, where those aged 35-44 years were more likely to be vaccine hesitant or resistant than accepting [27]. Consistent with other studies a higher level of education was associated with less likelihood of vaccine hesitancy [26,28]. Findings on socio-economic status are mixed, with individuals in the 2^nd^ tertile less likely to be hesitant and those in the 3^rd^ tertile being more hesitant, when compared to those in the 1^st^ tertile. However, with the exception of the rural pastoralist site, income was not associated with vaccine hesitancy. Other studies reported less vaccine hesitancy among higher income groups [13,27]. These findings illustrate variations among certain sub-groups in the population and hence the need for focussed messaging and campaigns aimed at specific target groups that are more likely to be vaccine hesitant.

Third, attitudes and level of compliance with other NPI COVID-19 measures appear to be associated with hesitancy. For example, those who reported difficulty in adhering to government regulations regarding COVID-19 prevention, reported wearing face masks less often now than when the pandemic started were more likely to be vaccine hesitant. Other studies have also reported an increased likelihood of vaccine hesitancy amongst those who are less adherent to measures put in place to control COVID-19 [29]. Reduced adherence could also be linked with reduced trust in institutions or government. Findings from a global survey found that those who reported having trust in their governments were less likely to be vaccine hesitant [28]. This finding highlights the need for public health messaging of vaccination to be holistic and include information about the value of other public health measures. As vaccine rollout in Kenya may take some time, NPI preventive measures must continue to avoid a resurgence of infections.

Fourth, individuals perceived risk of COVID-19 and its impact on their lives and livelihood also determined their level of hesitancy. For instance, those who had tested for COVID-19 previously, and those who had experienced substantial socio-economic impacts of COVID-19 were less likely to be hesitant. This finding is similar to findings in Saudi Arabia and France, where individuals with an increased perceived risk of COVID-19 were less likely to be vaccine hesitant [22,26]. This underlines the need to raise awareness about the consequences and risks of COVID-19 and to effectively communicate the value of vaccines.

Fifth, consistent with other studies, those who had concerns over COVID-19 vaccines’ side-effects and effectiveness were more likely to be vaccine hesitant [26,28–30]. This highlights the prevailing environment where there is heightened concern about the effectiveness and side effects of COVID-19 vaccines. Our findings emphasize the importance of a holistic, dynamic, transparent, and consistent public health messaging in improving vaccine hesitancy. Attention should be placed on building trust in the vaccine [31,32]. Additionally, reassurance of the capabilities of the regulatory bodies in ensuring safety and effectiveness should be emphasized [28]. This should be accompanied by open access, real time safety data at a national and regional level, and risk-based assessments that inform decision making.

This study focused on one dimension of access, namely acceptability. However, we recognize that other dimensions of access such as the availability of COVID-19 vaccines ensured through procurement and supply-side factors, and the affordability of the vaccines need to be considered over and above vaccine hesitancy as we aim to achieve herd-immunity, and the control of the pandemic. Further, this study has several limitations. First, the sample was drawn from existing Population Council cohorts whose households all have adolescents. Therefore, the sample is not representative of the four counties included in this study and the results are not generalizable to the full population. Second, the study was cross-sectional and reflects the level and determinants of vaccine hesitancy, as of February 2021. This was before the actual COVID-19 vaccine rollout in Kenya that started in March 2021. Conducting a longitudinal study would have provided more information on the change in vaccine hesitancy and its drivers, which could also inform the tailoring of messages over time. Third, there is also a need for qualitative studies to further explore the drivers and deterrents of COVID-19 vaccine uptake and the factors that may improve or compound COVID-19 vaccine acceptance. Despite these reported weaknesses, the study provides important insights on the COVID-19 vaccine hesitancy in certain locations of Kenya and provides implications to policymakers on possible avenues of improving vaccine hesitancy in Kenya.

## Conclusions

Our findings highlight that almost one year into the pandemic and about a month before vaccine rollout had begun in Kenya, vaccine hesitancy was quite high. As the current vaccine rollout plan will take time, there is also a need to promote holistic public health messaging to ensure that NPI interventions such as face masks, hand washing, physical distancing, and hand sanitizer use continue. These behaviours are tied to vaccine hesitancy and confidence, clearly linking the two and the need for cohesive messaging campaigns. We find significant variation by socio-demographics and perceived risk of COVID-19 infection and economic impacts; tailored messages may be required to reach those with different concerns, levels of education, and other factors. Lastly, there is a critical need for accurate and transparent information from trusted sources to combat misinformation particularly around vaccine side effects and effectiveness.

## Data Availability

The data presented in this study is available from the corresponding author on reasonable request

## Author Contributions

Conceptualization, K.A., T.A., E.B., M.H., J.P. and G.W.; formal analysis, S.O.; methodology, E.B., S.O., J.P., and D.M.; validation, T.A., M.H., G.W., K.A., and E.B.; investigation, J.P., D.M., T.A., and K.A.; data curation, D.M., J.P., and S.O.; writing—original draft preparation, S.O.; writing—review and editing, E.B., J.P., D.M., T.A., M.H., G.W., and K.A.; visualization, S.O., J.P., D.M., T.A., M.H., G.W., K.A., and E.B.; supervision, K.A., and E.B.; funding acquisition, K.A. All authors have read and agreed to the published version of the manuscript.

## Institutional Review Board Statement

The study was conducted according to the guidelines of the Declaration of Helsinki, and approved by the Institutional Review Board of Population Council (protocol code 936) and AMREF-Ethics and Scientific Review Committee (protocol P803/2020)

## Informed Consent Statement

Informed consent was obtained from all subjects involved in the study

## Data Availability Statement

The data presented in this study are available from the corresponding author on reasonable request

## REFERENCES

1. World Health Organization WHO Coronavirus Disease (COVID-19) Dashboard With Vaccination Data Available online: https://covid19.who.int/region/afro/country/ke (accessed on Jun 5, 2021).

2. Barasa, E.; Mothupi, M.C.; Guleid, F.; Nwosu, C.; Kabia, E.; Araba, D.; Orangi, S.; Muraya, K.; Gitaka, J.; Marsh, K. Health and socio-economic impacts of physical distancing for COVID-19 in Africa; 2020;

3. Barasa, E.; Kazungu, J.; Orangi, S.; Kabia, E.; Ogero, M.; Kasera, K. Assessing the Indirect Health Effects of the COVID-19 Pandemic in Kenya. CGD Work. Pap. 570 2021.

4. World Health Organization A Coordinated Global Research RoadMap: 2019 Novel Coronavirus; 2020;

5. World Health Organization Draft landscape and tracker of COVID-19 candidate vaccines Available online: https://www.who.int/publications/m/item/draft-landscape-of-covid-19-candidate-vaccines (accessed on Apr 25, 2021).

6. Anyiam-Osigwe, T. COVID-19 vaccines are now approved in some countries. What will it take to approve them for the rest of the world? Available online: https://www.gavi.org/vaccineswork/covid-19-vaccines-are-now-approved-some-countries-what-will-it-take-approve-them (accessed on Apr 25, 2021).

7. Kyobutungi, C. The ins and outs of Kenya’s COVID-19 vaccine rollout plan Available online: https://theconversation.com/the-ins-and-outs-of-kenyas-covid-19-vaccine-rollout-plan-156310 (accessed on Apr 26, 2021).

8. Hogan, A.B.; Winskill, P.; Watson, O.J.; Walker, P.G.; Whittaker, C.; Baguelin, M.; Haw, D.; Lochen, A.; Gaythorpe, K.A.M.; Imperial College COVID-19 Response Team; et al. Modelling the allocation and impact of a COVID-19 vaccine; 2020;

9. Schwarzinger, M.; Watson, V.; Arwidson, P.; Alla, F.; Luchini, S. COVID-19 vaccine hesitancy in a representative working-age population in France: a survey experiment based on vaccine characteristics. Lancet Public Heal. 2021, 6, e210–21, doi:10.1016/S2468-2667(21)00012-8.

10. SAGE Working Group on Vaccine Hesitancy *Report of the SAGE Working Group on Vaccine HEsitancy*; 2014;

11. Afolabi, A.A.; Ilesanmi, O.S. Dealing with vaccine hesitancy in Africa: The prospective COVID-19 vaccine context. Pan Afr. Med. J. 2021, 38, 1–7, doi:10.11604/pamj.2021.38.3.27401.

12. Nachega, J.B.; Sam-Agudu, N.A.; Masekela, R.; van der Zalm, M.M.; Nsanzimana, S.; Condo, J.; Ntoumi, F.; Rabie, H.; Kruger, M.; Wiysonge, C.S.; et al. Addressing challenges to rolling out COVID-19 vaccines in African countries. Lancet Glob. Heal. 2021, 0, doi:10.1016/s2214-109x(21)00097-8.

13. Dinga, J.N.; Sinda, L.K.; Titanji, V.P.K. Assessment of Vaccine Hesitancy to a Covid-19 Vaccine in Cameroonian Adults and Its Global Implication. Vaccines 2021, 9, 1–14, doi:10.3390/vaccines9020175.

14. Ditekemena, J.D.; Nkamba, D.M.; Mutwadi, A.; Mavoko, H.M.; Fodjo, J.N.S.; Luhata, C.; Obimpeh, M.; Van Hees, S.; Nachega, J.B.; Colebunders, R. Covid-19 vaccine acceptance in the Democratic Republic of Congo: A cross-sectional survey. Vaccines 2021, 9, 1–11, doi:10.3390/vaccines9020153.

15. World Health Organization Strategic Advisory Group of Experts *Report of the SAGE Working Group on Vaccine Hesitancy*; 2014;

16. Gallup *Wellcome Global Monitor-First Wave Findings*; 2019;

17. Abuya, T.; Austrian, K.; Isaac, A.; Kangwana, B.; Mbushi, F.; Muluve, E.; Mwanga, D.; Ngo, T.; Nzioki, M.; Ochako, R.; et al. Experiences among adults and adolescents during COVID-19 pandemic from four locations across Kenya-Study description; 2020;

18. Austrian, K.; Muthengi, E.; Mumah, J.; Soler-Hampejsek, E.; Kabiru, C.W.; Abuya, B.; Maluccio, J.A. The Adolescent Girls Initiative-Kenya (AGI-K): Study protocol. BMC Public Health 2016, 16, 210, doi:10.1186/s12889-016-2888-1.

19. Austian, K. Evaluation of the NISITU Program: a study to determine the effect of a gender attitudes and gender based violence program for adolescents in Nairobi, Kenya Available online: https://www.isrctn.com/ISRCTN67829143 (accessed on Apr 25, 2021).

20. Muthengi, E.; Austrian, K. Cluster randomized evaluation of the Nia Project: Study protocol. Reprod. Health 2018, 15, 218, doi:10.1186/s12978-018-0586-4.

21. Larson, H.J.; Jarrett, C.; Schulz, W.S.; Chaudhuri, M.; Zhou, Y.; Dube, E.; Schuster, M.; MacDonald, N.E.; Wilson, R.; Eskola, J.; et al. Measuring vaccine hesitancy: The development of a survey tool. Vaccine 2015, 33, 4165–4175, doi:10.1016/j.vaccine.2015.04.037.

22. Al-Mohaithef, M.; Padhi, B.K. Determinants of COVID-19 Vaccine Acceptance in Saudi Arabia: A Web-Based National Survey. J. Multidiscip. Healthc. 2020, 13, 1657–1663, doi:10.2147/JMDH.S276771.

23. Lazarus, J. V.; Ratzan, S.C.; Palayew, A.; Gostin, L.O.; Larson, H.J.; Rabin, K.; Kimball, S.; El-Mohandes, A. A global survey of potential acceptance of a COVID-19 vaccine. Nat. Med. 2020, 27, 225–228, doi:10.1038/s41591-020-1124-9.

24. Dzinamarira, T.; Nachipo, B.; Phiri, B.; Musuka, G. COVID-19 Vaccine Roll-Out in South Africa and Zimbabwe: Urgent Need to Address Community Preparedness, Fears and Hesitancy. Vaccines 2021, 9, 250, doi:10.3390/vaccines9030250.

25. IOL 52% of South Africans don’t want Covid vaccine, despite SA securing initial batch Available online: https://www.iol.co.za/personal-finance/insurance/52-of-south-africans-dont-want-covid-vaccine-despite-sa-securing-initial-batch-f45373f0-6199-46ab-8207-fb632706b8e5 (accessed on Apr 30, 2021).

26. Schwarzinger, M.; Watson, V.; Arwidson, P.; Alla, F.; Luchini, S. COVID-19 vaccine hesitancy in a representative working-age population in France: a survey experiment based on vaccine characteristics. Lancet Public Heal. 2021, 6, e210–e221, doi:10.1016/S2468-2667(21)00012-8.

27. Murphy, J.; Vallières, F.; Bentall, R.P.; Shevlin, M.; McBride, O.; Hartman, T.K.; McKay, R.; Bennett, K.; Mason, L.; Gibson-Miller, J.; et al. Psychological characteristics associated with COVID-19 vaccine hesitancy and resistance in Ireland and the United Kingdom. Nat. Commun. 2021, 12, 1–15, doi:10.1038/s41467-020-20226-9.

28. Lazarus, J. V.; Ratzan, S.C.; Palayew, A.; Gostin, L.O.; Larson, H.J.; Rabin, K.; Kimball, S.; El-Mohandes, A. A global survey of potential acceptance of a COVID-19 vaccine. Nat. Med. 2021, 27, 225–228, doi:10.1038/s41591-020-1124-9.

29. Edwards, B.; Biddle, N.; Gray, M.; Sollis, K. COVID-19 vaccine hesitancy and resistance: Correlates in a nationally representative longitudinal survey of the Australian population. PLoS One 2021, 16, e0248892, doi:10.1371/journal.pone.0248892.

30. Almaghaslah, D.; Alsayari, A.; Kandasamy, G.; Vasudevan, R. COVID-19 Vaccine Hesitancy among Young Adults in Saudi Arabia: A Cross-Sectional Web-Based Study. Vaccines 2021, 9, 330, doi:10.3390/vaccines9040330.

31. World Health Organization *Behavioural considerations for acceptance and uptake of COVID-19 vaccines*; Geneva, 2020;

32. Lawes-Wickwar, S.; Ghio, D.; Tang, M.Y.; Keyworth, C.; Stanescu, S.; Westbrook, J.; Jenkinson, E.; Kassianos, A.P.; Scanlan, D.; Garnett, N.; et al. A Rapid Systematic Review of Public Responses to Health Messages Encouraging Vaccination against Infectious Diseases in a Pandemic or Epidemic. Vaccines 2021, 9, 72, doi:10.3390/vaccines9020072.

