## Supplementary Table 1 for "Assessing the level and determinants of COVID-19 Vaccine Confidence in Kenya"

**Table S1**. Description of dependent and independent variables for the multivariate logistic regression

| Description | Categorization | Notes |
|---|---|---|
| **Dependent variable** | | |
| **Vaccine Hesitancy** | • Vaccine Hesitant (either somewhat likely, somewhat unlikely, very unlikely to accept the vaccine or don't know)<br>• Vaccine Accepting (Very likely to accept the vaccine) | |
| **Independent variables: Socio-demographic factors** | | |
| **County** | • Urban county (Nairobi/Kisumu)<br>• Rural county (Kilifi/Wajir) | |
| **Sex** | • Female<br>• Male | |
| **Age** | • 18-35 years<br>• 36-57 years<br>• 58+ years | |
| **Education** | • No schooling<br>• Pre-primary<br>• Primary education<br>• Secondary<br>• Tertiary | |
| **Marital status** | • Married<br>• Single | |
| **Total household size** | Continuous variable | |
| **Socio-economic status** | • Tertile 1 (Poorest)<br>• Tertile 2<br>• Tertile 3 (Wealthiest) | Principal component analysis was used to calculate socio-economic status based on (electricity, piped water source, reliable water source, livestock, and mobile phone ownership) and categorized into tertile. |
| **Independent variables: Individual influences, risk and perceptions** | | |
| **Perceived risk of getting COVID** | • Some risk (low risk, medium risk, high risk)<br>• No risk | |
| **Know someone in your family, neighbourhood or workplace who has been infected with COVID** | • Know someone who tested positive/suspected of having COVID<br>• Do not know someone who tested positive/suspected of having COVID | |

| | | |
|---|---|---|
| **Societal perception of having COVID** | 0. No perceived stigma<br>1. Perceived stigma | This was a composite variable of the following variables 1) People would stop talking to me 2) People would gossip about me 3) People I know would bring me the food I need 4) People I know would bring me the medicines I need 5) People in the community would treat me badly 6) After I recover from coronavirus, people in the community would still avoid me 7) After I recover from coronavirus, I would not be welcome back into my house by family 8) After I recover from coronavirus, I would not be welcome back at my place of work 9) After I recover from coronavirus, I would not be welcome back to my place of worship 10) After I recover from coronavirus, I would not be welcomed back to school |
| **Ease of following government regulations on COVID (q505)** | • Easy (Very easy to follow or somewhat easy to follow)<br>• Difficult (Somewhat difficult to follow and very difficult to follow) | |
| **Compared to the first few month of COVID, adherence to wearing of face masks over mouth and nose** | • More or about the same<br>• Less | |
| **Tested for Covid-19** | • Tested for COVID<br>• Never tested for COVID | |
| **Socio-economically impacted by COVID-19 measures** | • Socio-economically affected by measures<br>• Not socio-economically affected by measures | This is a composite variable of the following variables 1) skipped meals in the past 7 days because of not enough money or food 2) Complete loss of job as a result of COVID 3) Partial job loss as a result of COVID 4) Reduced healthcare access 5) economic status-making more or less than when the pandemic just began |
| **Independent variables: Context** | | |
| **Perceived measures taken by the community against COVID-19** | • Community in support of COVID-19 prevention measures<br>• Community does not support COVID-19 measures | This is a composite variable of the following variables: 1) Community taking steps to protect themselves and others from COVID-19 2) Community angry about social distancing measures due to COVID-19 3) Community work together to prevent and fight COVID-19 |
| **Government and media as trusted source of information** | • No<br>• Yes | This is a composite variable of the following variables 1) Government SMS 2) government television advertisements 3) chief/administrators 4) government radio advertisements 5) television programs/shows 6) govt television adverts 7) |

| | | |
|---|---|---|
| | | radio programs/shows 8) internet 9) posters/print adverts 10) social media |
| **Social Networks as trusted source of information** | • No<br>• Yes | This is a composite variable of the following variables: 1) Friends 2) acquaintances/neighbours 3) work colleagues |
| **Healthcare providers as trusted source of information** | • No<br>• Yes | This was a composite variable of the following variables: 1) Public health facility 2) private health facility 3) NGO provider 4) pharmacy 5) community health worker |
| **Community as trusted source of information** | • No<br>• Yes | This was a composite variable of the following variables: 1) public announcements with megaphones 2) church/mosque/sheikh/religious leader 3) community meetings/spaces |
| **Independent variables: Vaccination specific issues** | | |
| **Worry about side effects** | • No<br>• Yes | This was a composite variable combining: 1) vaccine trust 2) afraid of getting COVID after vaccination 3) worried about side effects |
| **Don't think the vaccine will be effective** | • No<br>• Yes | |
| **Too busy to get vaccinated** | • No<br>• Yes | |
| **Hard to access vaccination site** | • No<br>• Yes | |
| **Scared of needles** | • No<br>• Yes | |
| **For religious and cultural reasons** | • No<br>• Yes | |
